# Maternal Postpartum Stress Scale: Translation and validation study of the Chinese version

**DOI:** 10.1101/2023.07.31.23293354

**Authors:** Yanchi Wang, Qian Gao, Jin Liu, Feng Zhang, Xujuan Xu

## Abstract

**Objective:** This study aimed to translate the Maternal Postpartum Stress Scale (MPSS) into Chinese and validate its psychometric properties in postpartum women.

**Methods:** A total of 406 postpartum women were recruited from 6 hospitals in Nantong, Jiangsu, China. Cronbach’s α co-efficient, split-half reliability, and test-retest reliability were used to evaluate the reliability of the translated scale. Exploratory factor analysis (EFA) and confirmatory factor analysis (CFA) were used to evaluate the structural validity of the scale. The Edinburgh Postpartum Depression Scale (EPDS), the anxiety subscale of the Depression-Anxiety-Stress Scale (DASS-21) and the Perceived Stress Scale (PSS-10) were used as calibration scales to measure the correlation of MPSS.

**Results:** The Cronbach’s α co-efficient of the Chinese version of MPSS was 0.940 and the Cronbach’s α co-efficient of the three subscales ranged from 0.882 to 0.911. The split-half reliability was 0.825, and the test-retest reliability was 0.912. The content validity index of the scale (S-CVI) was 0.926. A total of three common factors were extracted from the EFA. The CFA validated the explored 3-factor structure, and the indicators fitted well (χ^2^/df = 2.167, comparative fit index = 0.918, tucker-lewis index=0.907, incremental fit index=0.919 and root mean square error of approximation = 0.075).

**Conclusion:** With superb reliability and validity, the Chinese version of MPSS can be used to assess postpartum stress in Chinese women, which is expected to lay a scientific basis for the early identification of postpartum women’s stress, thus providing a basis for the development of early individualized interventions. Equally importantly, with specific clinical value and practical significance for postpartum women’s physical and mental health, future replication studies of the Chinese version of MPSS should be carried out in diverse samples and clinical populations.

## 1. Introduction

For most parents, the birth of a child is a joyous event. However, the first year postpartum is also considered a critical transitional period, during which the mother can be emotional, accompanied by considerable distress [1]. Depression, anxiety and stress are the common postpartum psychological symptoms [2, 3], while nearly 67% of young mothers reported at least one mental health problem, and almost 40% had more than one [4]. Additionally, studies have shown that within one year after the birth of the first child, 14.2% of the mothers have depressive symptoms, 9.5% have moderate/severe anxiety symptoms, and 19.2% of the mothers have stress symptoms [5]. The incidence of postpartum depression, anxiety and stress is even higher in China, with 15.4% of the mothers having depressive symptoms, 15.9% having moderate/severe anxiety symptoms, and 41% having stress symptoms [6]. Therefore, an in-depth study of the causes of postpartum psychological changes and timely and effective interventions may benefit maternal postpartum mental health care.

Depression and anxiety during pregnancy and postpartum have been extensively researched over the past few decades because of their possible adverse effects on mothers’ and their offspring’s physical and mental health. Studies have shown significant associations between postpartum depression and internalizing and externalizing psychopathologies in children [7], while postpartum anxiety harms children’s temperament, sleep, and cognitive development [8]. However, postpartum stress, compared to postpartum anxiety and postpartum depression, has not been received widespread attention.

Postpartum stress is a particular type of stress that is conceptualized as an adverse psychological reaction to numerous obligations to raise a child, the existence of which is a rule rather than an exception [9]. Factors including infant care demands and changing social role expectations may be associated with increased stress of the mother. There is growing evidence that postpartum stress, one of the most common psychiatric disorders in postpartum woman, is associated with a high disease burden. Personally, a postpartum woman with stress usually suffers from intense, excessive, and persistent concerns and fears about daily situations [10, 11]. Postpartum women who cannot cope positively may have doubts about themselves and deviating self-evaluation. In this case, postpartum depression is prone to occur, which can even develop into serious postpartum mental illnesses such as postpartum depression.[12]. It can be seen that being more common than postpartum depression, postpartum stress affects a considerable proportion of mothers [2]. Postpartum stress is almost everywhere, which is worrying, especially during the COVID-19 pandemic, when it negatively impacts individuals’ quality of life, the growth and development of infants, family relations, and social functioning, as well as causes severe mental health problems [13, 14].Taken together, all these findings highlight the importance of understanding and screening postpartum women with high stress.

Postpartum stress is often under-identified and unde-diagnosed because it is frequently comorbid or co-occurs with numerous other psychiatric disorders, such as anxiety disorders and major depression [15, 16]. In turn, these diseases go unrecognized and underdiagnosed, making treatment difficult, resulting in lower remission rates and worse prognoses. Therefore, it is urgent to develop accessible and reliable instruments to screen and diagnose postpartum stress. Although structured interviews have been widely used to diagnose postpartum stress, these interviews are sometimes time-consuming, requiring some trained professionals to perform [17]. Moreover, as the first step in screening and early detection, self-report measures are a good, efficient and easy way to screen for postpartum stress. Currently, some scales for assessing postpartum stress have proven reliable and valid. However, the tools that evaluate postpartum stress are limited. Furthermore, there are problems such as redundant questionnaire items, lack of specificity, and measurement time being too early or too late.

There are few screening tools in China to assess postpartum stress, especially those that are culturally consistent with the Chinese culture, leading to less research on postpartum stress and related management in China. The postpartum stress evaluation is still a relatively new concept in the field of obstetric care in China. Although Hung Postpartum Stress Scale (HPSS) [6] developed by Hung et al. in 1993 was widely used in Taiwan, China, it was only applicable to the early postpartum period (within 6 weeks after delivery). Besides, few or no thoroughly evaluated features of the measurements were found. Moreover, the scale contained too many items and obsolete content. Therefore, there is still insufficient evidence to support HPSS as a specific parental stress measurement tool in China. It is, therefore, essential to develop effective, reliable, and specific assessment tools in a timely and accurate manner and to respond to and manage the symptoms of postpartum stress effectively.

Research on postpartum stress has increased significantly in recent decades. In a scoping review, there are presently 15 instruments, some of which are relevant psychometric information used to measure parental stress for parents with young children [18]. However, the amount of information about the psychometric properties of these tools is minimal. To assess the stress of women during postpartum maternity leave, Sandra et al. [19] developed the Maternal Postpartum Stress Scale (MPSS) in 2021. MPSS is a self-assessment questionnaire including 22 items. The results of exploratory factor analysis showed that three subscales (personal needs and fatigue (9 items), infant nurturing (7 items), body changes and sexuality (6 items)) was acceptable, and demonstrated good internal consistency (Cronbach’s alpha = 0.88). The introduction of MPSS has enriched the evaluation tools for postpartum stress. By understanding postpartum women’s stress at different times, MPSS is expected to be utilized to identify people at high risk of postpartum stress, which may help with individualized interventions. To the best of our knowledge, except for the original English version, the MPSS has not been validated in other ethnic groups.

In this study, to verify the psychometric properties of the MPSS in Chinese postpartum women, the original English version of MPSS was first translated into Chinese and culturally modified. Then, a total of 406 women were recruited to evaluate the reliability and validity of the translated MPSS.

### 2. Methods

#### 2.1 Participants and procedures

This cross-sectional study was conducted in the postpartum outpatient clinics of six hospitals in Nantong, Jiangsu Province, China from August to November 2022. According to the rule of thumb for sample size [20], a total of 425 postpartum women were recruited in the survey, of which 406 participants completed the MPSS in Chinese version (the response rate is 95.53%). Subjects were divided into two groups using a split-sampling random procedure. Group one, which included 200 participants, was used for exploratory factor analysis (EFA), while group two, which included 206 participants, was used for confirmatory factor analysis (CFA) [21]. During the survey, standardized instructions were presented to the participating postpartum women with a detailed explanation of the content and purpose of the investigation. All postpartum women were informed that participation was voluntary and that their responses were confidential. The survey lasted about 20-30 min. The selected postpartum women could withdraw at any time if they feel uncomfortable. Inclusion criteria: (1)≥18 years old; (2) single pregnancy, full-term live birth; (3)There were no malformations or serious complications in neonates. (4)Have reading comprehension ability, average cognition, and complete language expression ability; (5) Volunteer to participate in this study. Exclusion criteria: (1) Suffer from physical or mental disorders.

Prior to the survey, written informed consent was also obtained from all participants. Ethical approval for this study was obtained from the Research Ethics Committee of the corresponding author’s institution (approval number: 2022-K50-01).

#### 2.2 Measures

A questionnaire consisting of sociodemographic questions and 22 items of the translated MPSS was constructed. Besides, several classical scales including the Edinburgh Postnatal Depression Scale (EPDS) [22], the Depression Anxiety Stress Scale-21 (DASS-21) [23] were applied to evaluate divergent validity and the 10-item Perceived Stress Scale (PSS-10) [24] was applied to evaluate convergent validity.

##### 2.2.1 Maternal Postpartum Stress Scale (MPSS)

MPSS [19] is a reliable and valid instrument measuring self-reporting postpartum Stress among postpartum women. This scale contains 22 items measuring three subscales of postpartum stress: personal needs and fatigue (9 items), infant nurturing (7 items), body changes and sexuality (6 items). Each item of the scale is scored on a 4-point Likert scoring ranging from 0 (not at all) to 4 (completely). The higher the overall MPSS score, the greater the postpartum stress.

##### 2.2.2 Translation of the MPSS

After obtaining permission from the original authors, this scale was translated into Chinese using the classical “backward and forward” procedure following a modified Brislin translation model [25]. Two independent, bilingual Chinese-native psychologists performed the forward translation of items from English into Chinese. After all authors’ and translators’ discussion and revision, a consensus forward Chinese version of the instrument was developed. Subsequently, the reconciled forward version of the scale was back-translated into English by two independent bilingual psychologists blinded to the original English version. In the third step, a semantic comparison of the original and post-translated versions of the scale was carried out according to the cross-cultural adaptation guidelines of the scale [26], which was reviewed by a panel of 15 sociologists, psychologists, nursing education experts, clinical medical experts, and obstetric nursing experts. Subsequently, 30 postpartum women were interviewed cognitively to investigate the comprehensibility of MPSS Chinese translations with appropriate modifications [27]. The final version of MPSS is presented in **Supplementary Table 1**. The Item Content Validity Index (I-CVI) ranged from 0.88-1.00. After evaluation by 8 experts, the average scale content effectiveness index (S-CVI) turned out to be 0.926 [28]. Cronbach’s α coefficient of internal consistency of the scale in this study was 0.940.

##### 2.2.3 Edinburgh Postnatal Depression Scale (EPDS)

The Chinese version of the Edinburgh Postnatal Depression Scale (EPDS) [22] was used to assess the postpartum depression status of postpartum women, which consists of 10 items. Each item response is divided into 4 levels, reflecting the severity of symptoms, from “never” to “always” with scores ranging from 0 to 3, respectively. The scores of the 10 items were summed to obtain the total individual score, which ranged from 0 to 30. We used 10 as the cut-off point.

##### 2.2.4 Depression Anxiety Stress Scale-21 (DASS-21)

The Depression, Anxiety, and Stress Scale−21 (DASS-21) [23] were used to measure depressive, anxiety, and stress symptoms among participants. DASS-21 comprises 21 self-reported items with three subscales: depression, anxiety, and stress. Over the past week, respondents were asked to rate each item from 0 (did not apply to me at all) to 3 (applied to me very much). For compatibility with the DASS-42, the DASS-21 score is multiplied by 2. Psychometric properties of the scale in assessing depression, anxiety, and stress were well established, including in pregnant and postpartum women. This study should focus on the anxiety subscale [29].

##### 2.2.5 10-item Perceived Stress Scale (PSS-10)

Stress severity was assessed by the validated Chinese version of the 10-item Perceived Stress Scale (PSS-10) [24], which has been widely used in clinical research that measures general stress levels. Each item is rated on a 5-point scale, ranging from 0 (*never*) to 4 (*very often*), with a total score ranging from 0 to 40, whereby a higher score indicates a higher level of stress [30].

#### 2.3 Statistical analyses

The EFA and CFA were used to explore and validate the potential factor structure of the scale, respectively. The 406 participants were divided into two groups according to the principle of randomization, with 200 and 206 participants assigned to EFA and CFA tests. In EFA, the scale is suitable for factor analysis only if KMO > 0.6 and the Bartlett spherical test was statistically significant (*P* < 0.05) [31]. Amos software (23.0) was used to verify the model structure’s consistency with the exploration factors’ structure. These indices were used to examine the goodness of model fit: the comparative fit index (CFI). When Chi-square/df (χ^2^/df)<3.000, Root mean square residual(RMR)<0.050, and root-mean-square error of approximation (RMSER) < 0.080, and the model fits well if the value increment fitting index (IFI), Tucker Lewis index (TLI), and comparative fitting index (CFI) exceed 0.900 [32]. Reliability for internal consistency of the scale was determined based on Cronbach’s alpha coefficient. The Cronbach’s alpha coefficient exceeding 0.7 was considered acceptable [33]. The two-sided chi-squared test was used to statistically evaluate differences in demographic characteristics between the EFA and CFA groups. The Student’s t-test was used to statistically evaluate differences in total MPSS and subscale scores for postpartum women in different statuses. All data were analyzed using SPSS 25.0 and AMOS 24.0, with a significant α threshold of 0.05

(two-tailed).

### Results

#### Demographic information

The characteristics of the subjects in EFA (n=200) and CFA (n=206) in this study are summed up in **Table 1**. All the variables were comparable between the two groups (*P* >0.05).

**Table 1.**
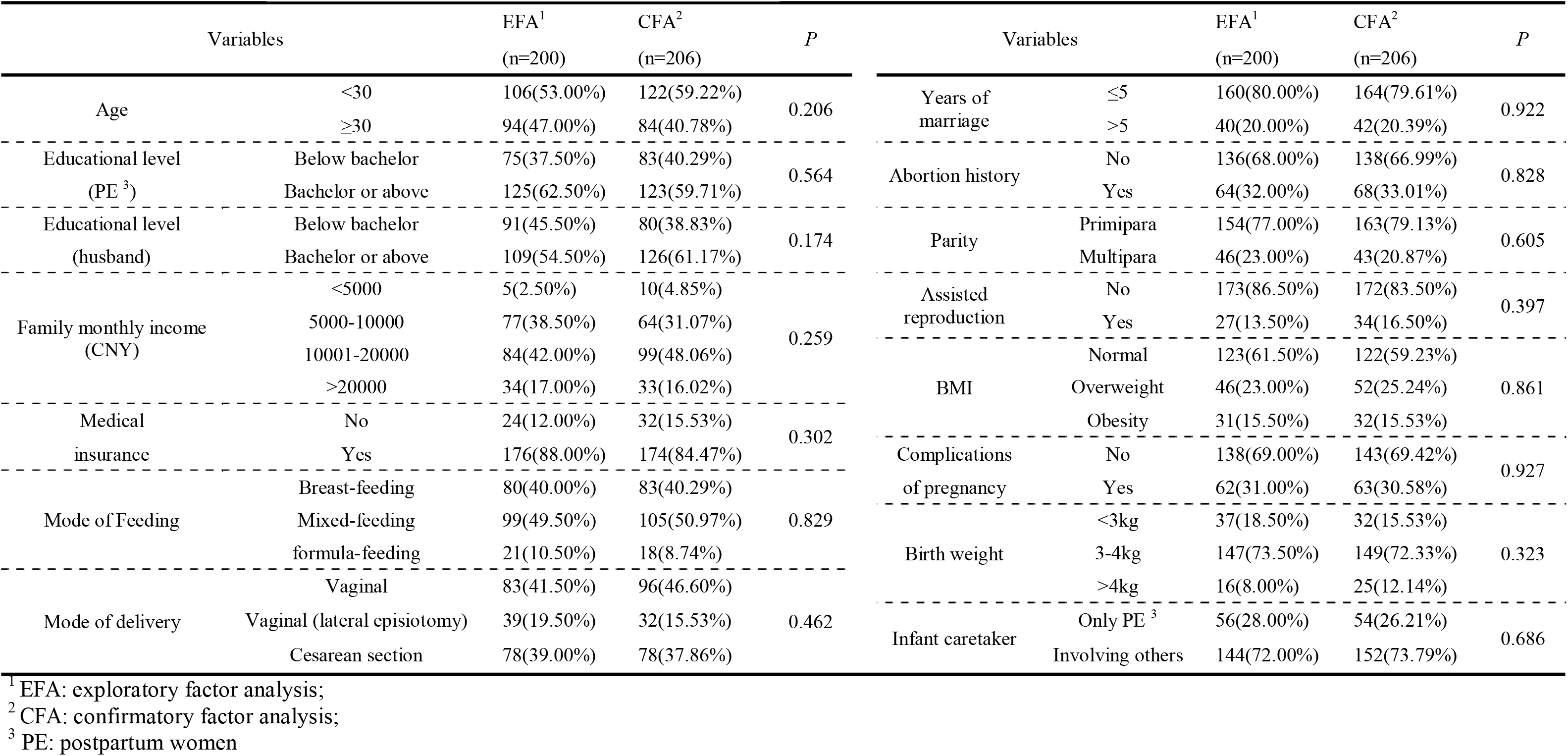
Characteristics of the subjects enrolled in this study.

**Table 2.** The results of the discriminant validity analysis using the Pearson’ s correlation coefficient.

|  | A1 | A2 | A3 | A4 | A5 | A6 | A7 | A8 | A9 | A10 | A11 | A12 | A13 | A14 | A15 | A16 | A17 | A18 | A19 | A20 | A21 | A22 | Total score |
| --- | --- | --- | --- | --- | --- | --- | --- | --- | --- | --- | --- | --- | --- | --- | --- | --- | --- | --- | --- | --- | --- | --- | --- |
| A1 | 1 | .623** | .513** | .650** | .639** | .586** | .380** | .469** | .390** | .353** | .288** | .255** | .283** | .371** | .305** | .271** | .291** | .381** | .468** | .412** | .320** | .399** | .687** |
| A2 | .623** | 1 | .613** | .585** | .587** | .496** | .445** | .494** | .485** | .356** | .271** | .252** | .326** | .301** | .270** | .260** | .262** | .440** | .501** | .369** | .407** | .451** | .679** |
| A3 | .513** | .613** | 1 | .577** | .584** | .490** | .466** | .446** | .442** | .352** | .208** | .341** | .312** | .351** | .288** | .246** | .285** | .345** | .443** | .377** | .322** | .355** | .653** |
| A4 | .650** | .585** | .577** | 1 | .839** | .735** | .456** | .465** | .391** | .370** | .275** | .338** | .362** | .388** | .349** | .294** | .312** | .415** | .479** | .446** | .337** | .408** | .743** |
| A5 | .639** | .587** | .584** | .839** | 1 | .759** | .473** | .466** | .417** | .390** | .267** | .338** | .314** | .362** | .324** | .296** | .298** | .417** | .477** | .420** | .361** | .411** | .741** |
| A6 | .586** | .496** | .490** | .735** | .759** | 1 | .548** | .456** | .426** | .406** | .295** | .348** | .334** | .355** | .319** | .308** | .334** | .355** | .483** | .435** | .412** | .431** | .726** |
| A7 | .380** | .445** | .466** | .456** | .473** | .548** | 1 | .465** | .437** | .385** | .266** | .230** | .290** | .299** | .240** | .247** | .239** | .337** | .404** | .328** | .381** | .343** | .595** |
| A8 | .469** | .494** | .446** | .465** | .466** | .456** | .465** | 1 | .699** | .556** | .507** | .479** | .357** | .373** | .353** | .312** | .320** | .591** | .746** | .451** | .594** | .578** | .752** |
| A9 | .390** | .485** | .442** | .391** | .417** | .426** | .437** | .699** | 1 | .532** | .386** | .441** | .275** | .334** | .322** | .274** | .281** | .527** | .596** | .367** | .568** | .524** | .678** |
| A10 | .353** | .356** | .352** | .370** | .390** | .406** | .385** | .556** | .532** | 1 | .367** | .452** | .350** | .374** | .362** | .296** | .266** | .544** | .549** | .410** | .554** | .538** | .653** |
| A11 | .288** | .271** | .208** | .275** | .267** | .295** | .266** | .507** | .386** | .367** | 1 | .489** | .290** | .309** | .304** | .348** | .218** | .415** | .484** | .297** | .433** | .427** | .534** |
| A12 | .255** | .252** | .341** | .338** | .338** | .348** | .230** | .479** | .441** | .452** | .489** | 1 | .320** | .364** | .349** | .331** | .241** | .359** | .512** | .334** | .413** | .467** | .572** |
| A13 | .283** | .326** | .312** | .362** | .314** | .334** | .290** | .357** | .275** | .350** | .290** | .320** | 1 | .611** | .591** | .528** | .461** | .452** | .466** | .509** | .438** | .400** | .615** |
| A14 | .371** | .301** | .351** | .388** | .362** | .355** | .299** | .373** | .334** | .374** | .309** | .364** | .611** | 1 | .844** | .572** | .526** | .449** | .431** | .603** | .406** | .411** | .672** |
| A15 | .305** | .270** | .288** | .349** | .324** | .319** | .240** | .353** | .322** | .362** | .304** | .349** | .591** | .844** | 1 | .604** | .480** | .466** | .422** | .526** | .447** | .425** | .637** |
| A16 | .271** | .260** | .246** | .294** | .296** | .308** | .247** | .312** | .274** | .296** | .348** | .331** | .528** | .572** | .604** | 1 | .374** | .398** | .383** | .419** | .434** | .415** | .565** |
| A17 | .291** | .262** | .285** | .312** | .298** | .334** | .239** | .320** | .281** | .266** | .218** | .241** | .461** | .526** | .480** | .374** | 1 | .455** | .393** | .698** | .410** | .367** | .574** |
| A18 | .381** | .440** | .345** | .415** | .417** | .355** | .337** | .591** | .527** | .544** | .415** | .359** | .452** | .449** | .466** | .398** | .455** | 1 | .671** | .531** | .633** | .570** | .718** |
| A19 | .468** | .501** | .443** | .479** | .477** | .483** | .404** | .746** | .596** | .549** | .484** | .512** | .466** | .431** | .422** | .383** | .393** | .671** | 1 | .519** | .648** | .592** | .785** |
| A20 | .412** | .369** | .377** | .446** | .420** | .435** | .328** | .451** | .367** | .410** | .297** | .334** | .509** | .603** | .526** | .419** | .698** | .531** | .519** | 1 | .468** | .459** | .704** |
| A21 | .320** | .407** | .322** | .337** | .361** | .412** | .381** | .594** | .568** | .554** | .433** | .413** | .438** | .406** | .447** | .434** | .410** | .633** | .648** | .468** | 1 | .674** | .703** |
| A22 | .399** | .451** | .355** | .408** | .411** | .431** | .343** | .578** | .524** | .538** | .427** | .467** | .400** | .411** | .425** | .415** | .367** | .570** | .592** | .459** | .674** | 1 | .706** |
| Total score | .687** | .679** | .653** | .743** | .741** | .726** | .595** | .752** | .678** | .653** | .534** | .572** | .615** | .672** | .637** | .565** | .574** | .718** | .785** | .704** | .703** | .706** | 1 |

Among the total 406 participants, the median score of the MPSS was 15, while the median scores of the three subscales of the MPSS were 5 (personal needs and fatigue), 6 (infant nurturing), and 3 (body changes and sexuality), respectively. Details of the number of participants in each score group were showed in **Figure 1**.

**Figure 1.**
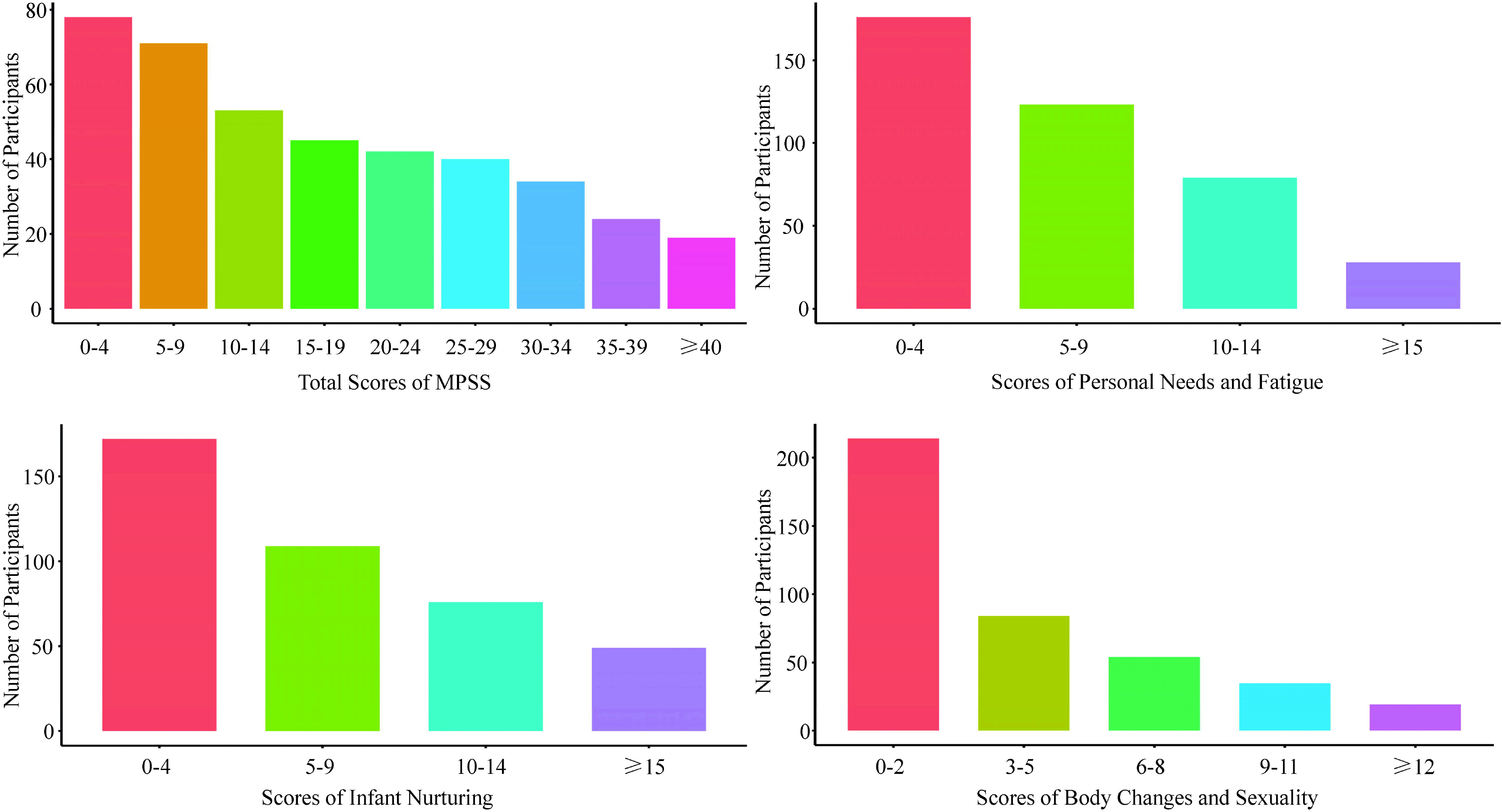
Number of participants in each MPSS and subscale groups.

#### Item analysis

The Chinese version of MPSS of the 406 survey respondents was sorted by their score. The top 27 percent of those who scored were grouped into high stress group, while those who scored the bottom 27 percent were divided into low stress group. Furthermore, the average score for each item in both groups was calculated. Two-tailed independent samples t-test showed there was a significant difference between the items in the two groups (*p*<0.05) (**Supplementary Table 2)**. The correlation analysis results (**Table2**) revealed that the correlation coefficients among all items in the scale were > 0.4, indicating that each item had good correlation with the dimension, and no item was deleted. The results of discrete trend method (**Supplementary Table 3**) showed that the coefficient of variation of each item was >0.80. All items met the inclusion criteria.

#### Content Validity

As shown in **Supplementary Table 4**, the CVI of each item in the MPSS was 0.88 to 1, indicated that the items were comprehensible to the target users [34], and the S-CVI of the MPSS was 0.926.

#### Reliability analysis

The Cronbach’s α coefficient of the total MPSS is 0.940, while the Cronbach’s α coefficients of the three subscales are 0.903, 0.911 and 0.882, respectively. The half-test reliability is 0.825 and the retest reliability is 0.912 [35], indicating good reliability (**Supplementary Table 5**). As shown in **Supplementary Table 6**, the correlation coefficient between the 22 items of the questionnaire and the total score, and the Cronbach alpha coefficients after removing one item from the questionnaire, was lower than the Cronbach alpha coefficient of 0.90 before the removal.

#### Validity analysis

##### Exploratory factor analysis

In this study, KMO is 0.910, and Bartlett’s spherical test was statistically significant χ^2^ =3511.060, *P* < 0.01), indicating that the translated MPSS was suitable for factor structure analysis. A total of three factors with eigenvalues > 1 were extracted and a total of 65.716% of the data discrepancies were explained. Furthermore, the results of EFA showed that each factor load was > 0.4. Therefore, all items were retained. The specific results can be found in **Table 3**.

**Table 3.** Exploratory factor analysis of the MPSS in Chinese version (n=200)

| Items | factor 1 | factor 2 | factor 3 |
| --- | --- | --- | --- |
| A21 Impossibility to complain to someone | 0.805 |  |  |
| A8 Adjustment to frequent wake-ups | 0.759 |  |  |
| A9 Baby's irregular patterns of daily sleep | 0.754 |  |  |
| A22 Loneliness at home with the baby | 0.737 |  |  |
| A18 Lack of time for socializing with friends | 0.731 |  |  |
| A10 My fatigue and exhaustion | 0.669 |  |  |
| A19 Lack of time for myself | 0.650 |  |  |
| A12 Lack of help with the baby and household chores | 0.604 |  |  |
| A11The amount of the household chores | 0.571 |  |  |
| A5 Baby's health problems |  | 0.827 |  |
| A4 Baby's development |  | 0.823 |  |
| A6 Recognizing the baby's needs |  | 0.763 |  |
| A3 Insufficient milk supply when breastfeeding |  | 0.759 |  |
| A2 Baby's irregular feeding pattern |  | 0.743 |  |
| A1 Choosing the appropriate way of feeding the baby |  | 0.625 |  |
| A7 Impossibility to soothe a crying or upset baby |  | 0.560 |  |
| A15 Insufficient enjoyment in sexual intercourse |  |  | 0.856 |
| A14 Insufficient frequency of sexual intercourse |  |  | 0.833 |
| A13 Being uncertain when to resume intercourse after childbirth |  |  | 0.737 |
| A16 The thought that my partner finds me unattractive |  |  | 0.713 |
| A20 Physical appearance after childbirth |  |  | 0.704 |
| A17 The impossibility to return to the pre-pregnancy weight |  |  | 0.644 |
| Eigenvalues | 10.486 | 2.153 | 1.595 |
| Cumulative variance contribution rate (%) | 23.439 | 45.415 | 64.699 |

##### Confirmatory factor analysis

Confirmatory factor analysis was used in this study to indicate whether the structure of the questionnaire was consistent with the theoretical structure of the original questionnaire. The results after model modification are shown in **Figure 2**. Confirmatory factor analysis was used to validate whether the relationship between each item and the factors was consistent with the hypothesis. In this study, the validation results demonstrated that the fittings were good. The values of the indicators were: χ^2^/df = 2.167(<3), CFI= 0.918(>0.9), IFI=0.919(>0.9). TLI=0.907(>0.9), and RMSEA=0.075(<0.08) (**Supplementary Table 7**).

**Figure 2.**
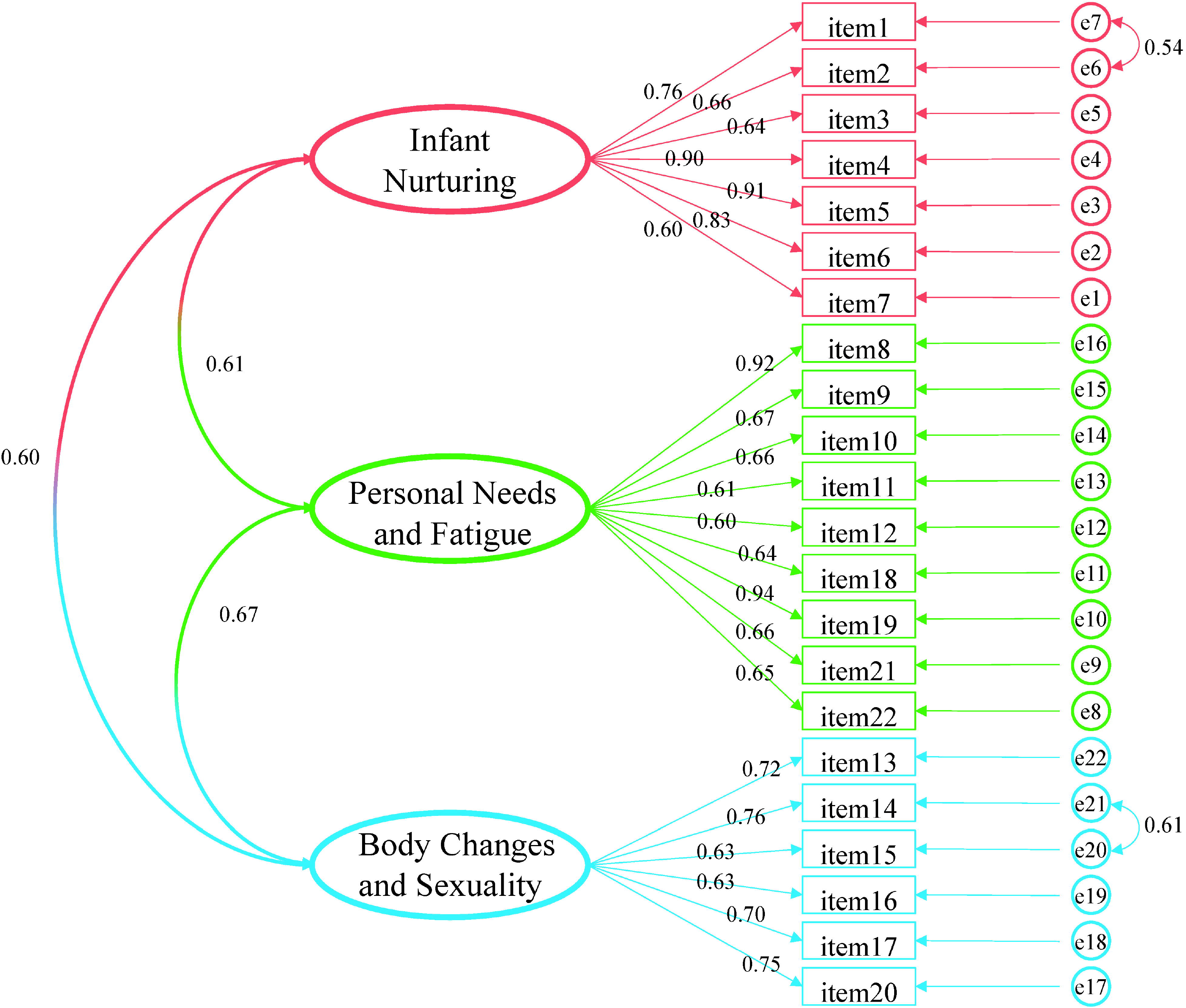
Confirmatory factor analysis model of the Chinese version of MPSS.

##### Calibration correlation validity

This study used EPDS, DASS-21 anxiety subscale and PSS-10 as calibration. As shown in **Figure 3**, the EPDS test results showed that there were 75 women (18.47%) with depression. The MPSS score of depressed women was significantly higher than that of non-depressed women, with a statistically significant difference (*P*<0.05). The scores were higher on all subscales than in non-depressed women (*P*<0.05). The results of the anxiety subscale in DASS-21 showed that 68 women (16.75%) were in an anxious state. The MPSS score of anxious women was significantly higher than that of non-anxious women (*P*<0.05). All subscales had higher scores than those without anxiety (*P* < 0.05). The PSS-10 results showed that postpartum women with high-stress perception scores had significantly higher MPSS scores than those with low-stress perception scores (*P*<0.05). Moreover, the MPSS subscales also showed the same trend (*P* < 0.05). The Pearson correlation coefficients between the revised Chinese version of MPSS and EPDS, the anxiety subscale in DASS-21 and the PSS-10 were 0.502, 0.404 and 0.476 (*P* < 0.01), respectively. The correlation coefficients between each subscale of MSPP and EPDS, the anxiety dimension in DASS-21 and the PSS-10 are shown in **Figure 4**.

**Figure 3.**
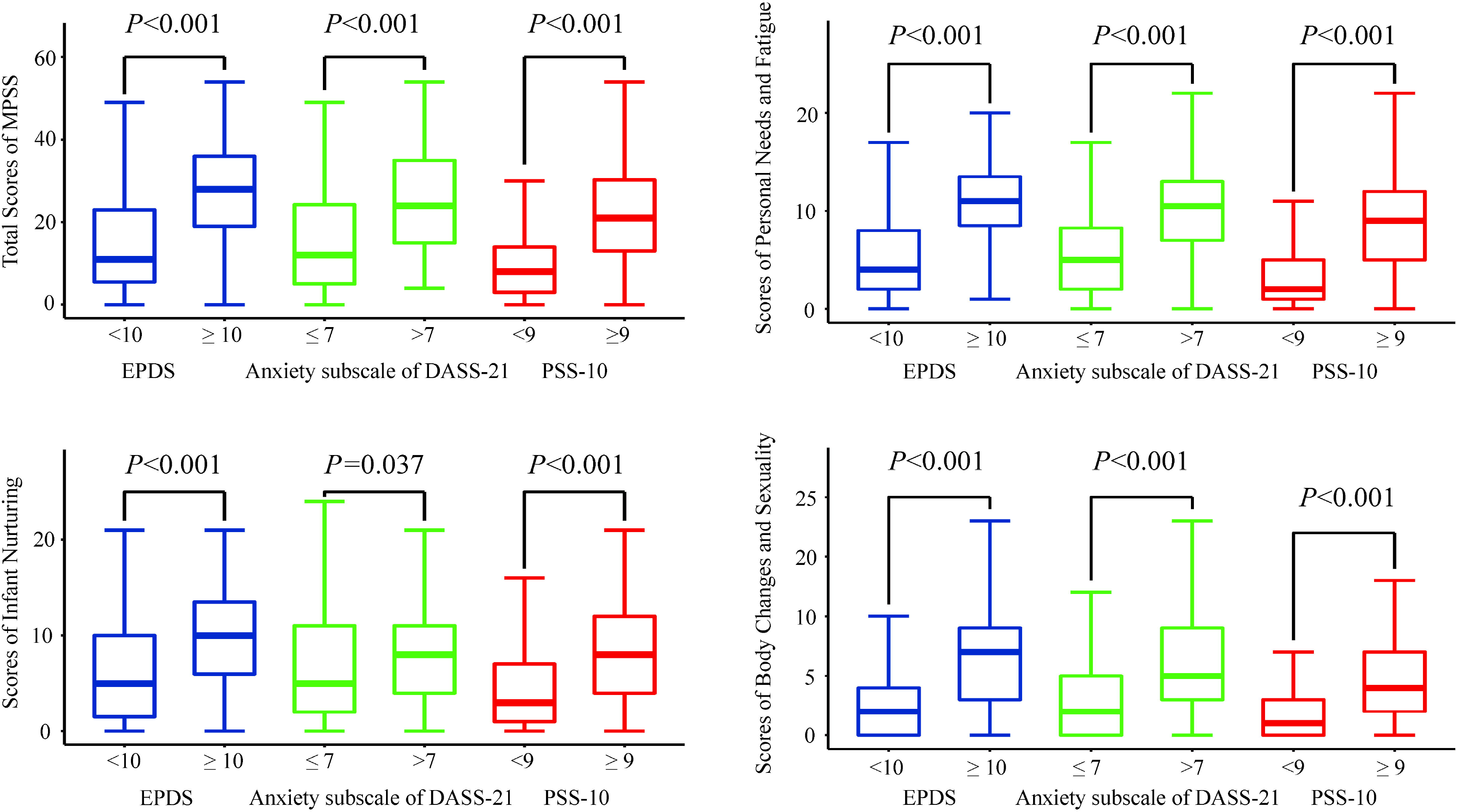
The Scores of MPSS for Maternity in Different Status.

**Figure 4.**
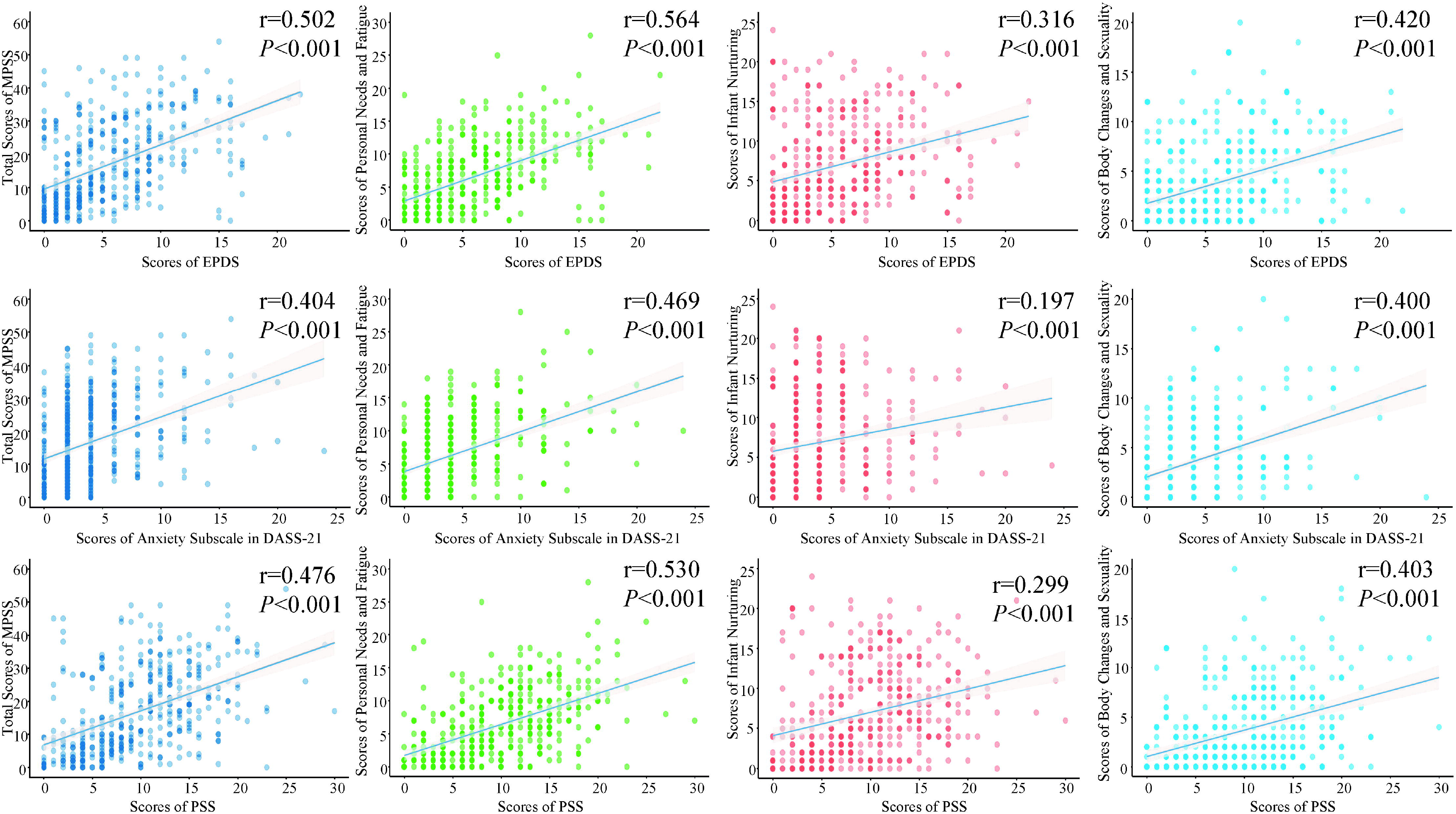
Correlation between EPDS, DASS-21, PSS-10, and the Chinese version of MPSS.

##### The correlation between the total items of MPSS and each subscale

Quantitatively, each subscale of postpartum stress has an impact on the total score of postpartum stress: personal needs and fatigue (r=0.890), infant nurturing (r=0.870), body changes and sexuality (r=0.796). Personal needs and fatigue were most strongly associated with infant nurturing (r=0.6440) (**Figure 5**).

**Figure 5.**
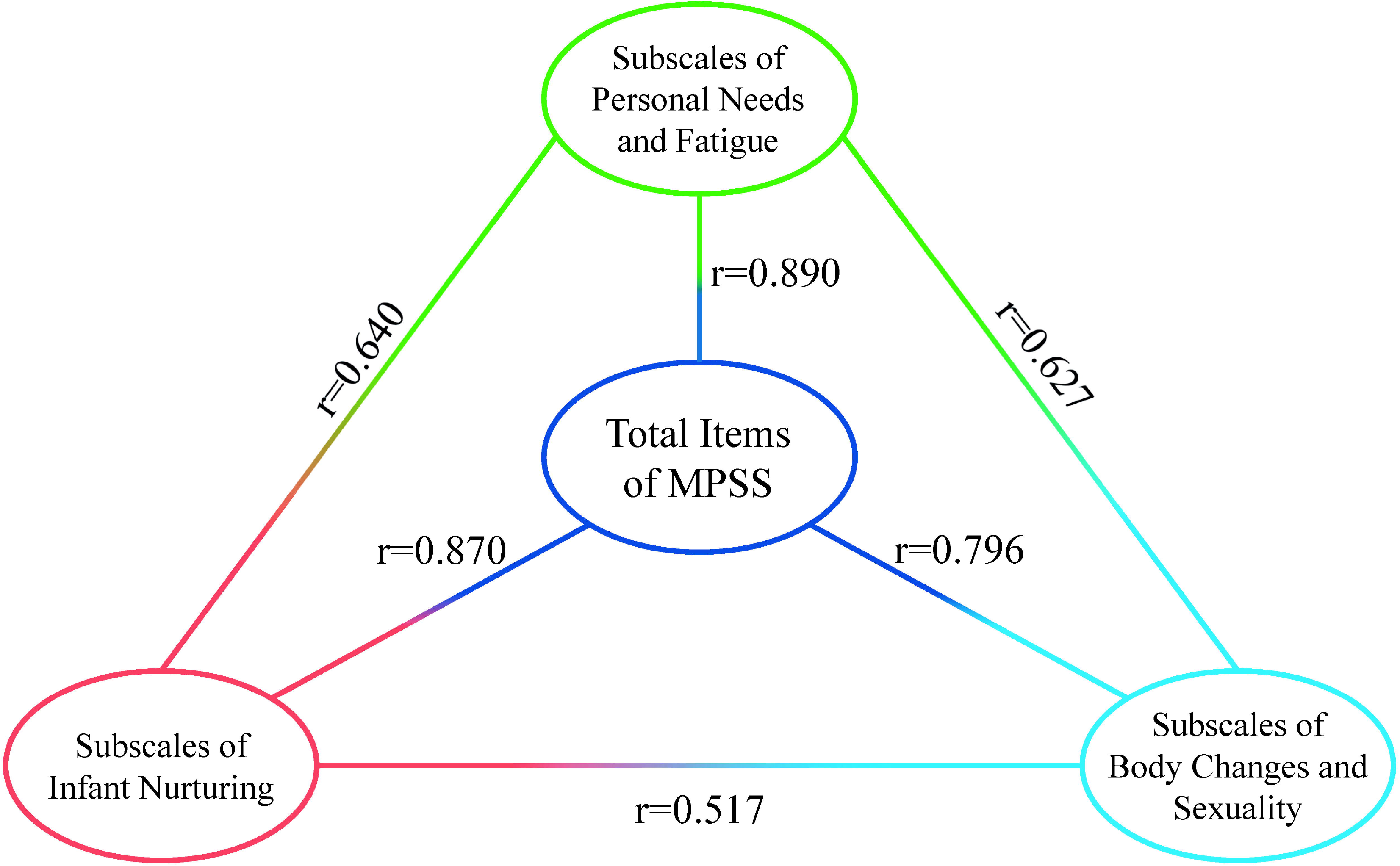
Correlation between total items of MPSS and each subscale.

## Discussion

Maternal postpartum stress represents an essential aspect of women’s mental health after childbirth [36]. Hence, there are indeed scales to assess postpartum stress. However, lacking specificity, most of them are universal scales. Even if they are specific, they still have shortcomings such as too early or too late measurement time and too much content. [1, 37, 38] At present, there is no specific tool that can measure postpartum stress from multiple perspectives. When the MPSS scale was developed in Croatia, it was applicable to women one year after childbirth. By contacting Sandra et al. and we learned that women in Croatia are entitled to paid leave in the first year after giving birth. Therefore, this scale can also measure stress of postpartum women during maternity leave in China. In addition, applying the MPSS scale in postpartum women in China can provide clinicians with tools to quickly identify stress in postpartum women, which can significantly promote the effective and accurate management of health psychology in postpartum women

In this study, we followed strict guidelines for cross-cultural adaptation of MPSS and tested the psychometric properties of MPSS in the Chinese population. The MPSS was finally formed through the cultural modification of expert letter consultation and cognitive interview. Expert group members are highly professional and include obstetricians, gynecologists, nursing education specialists, sociologists, pediatricians, psychologists, statisticians, and clinicians. Expert group members have strong comprehensive strength and comprehensive involvement in the field, which promotes the objectivity and professionalism of scale evaluation. A formal survey of 406 postpartum women showed that the Chinese version of MPSS had fabulous reliability and validity. The Chinese version of MPSS contains 3 subscales and 22 items in total. The scale items are moderate with the filling time less than 5 min. The contents of the items are clear and easy to understand. The Chinese version of MPSS evaluates postpartum stress from multiple dimensions, including personal needs and fatigue, infant nurturing, body changes and sexuality, which is expected to provide data reference for medical staff to better evaluate postpartum stress, and develop individualized intervention programs for postpartum stress to improve postpartum stress.

The S-CVI of the Chinese version of MPSS was 0.926 > 0.900, and the I-CVI was 0.88-1.00 > 0.800. After cognitive interview, respondents systematically analyzed each scale item in depth. The understanding and acceptance of each scale item were improved by optimizing the scale to adapt it to the Chinese cultural context. It indicates that the content validity of the scale is good. In this study, saturated with three factors explaining 65.716% of the items’ variance. However, in the original scale, saturated with three factors explaining 36.1% of the items’ variance. In this study, both exploratory factor analysis and confirmatory factor analysis suggested that the scale structure was stable. In addition, the EPDS and the anxiety subscale of DASS-21 were applied to evaluate divergent validity and the PSS-10 was applied to evaluate convergent validity. The correlation coefficients between MPSS in the Chinese version and EPDS, anxiety subscale in DASS-21and PSS-10 were 0.502, 0.404 and 0.476 (*P* < 0.01), respectively. The total scale and all subscales correlated moderately with general perceived stress. The Personal needs and fatigue subscale was in the highest correlation with general perceived stress. Other studies support this finding in a way that mothers in the postpartum period reported a lack of time for themselves and exhaustion [39]. Also, the total scale and all subscales are associated with depression and anxiety. This is consistent with previous research which found that mothers who experience more stress are at risk for mental health issues [40]. However, it should be noted that the convergent validity was only moderate, with the MPSS and general stress scale sharing only 47.6% of the variance. Similar patterns were seen in other postnatal stresses, suggesting a small overlap between specific and general postnatal stress [41–43].Reliability reflects the internal consistency and stability of the results measured by the measuring tool. In this study, the Cronbach’s α coefficient of the total scale was 0.940, while the Cronbach’s α coefficients of the three subscales are 0.903, 0.911 and 0.882, respectively. The Cronbach’s α coefficient of the original scale was 0.880, while the Cronbach’s α coefficient of the three subscales was 0.850, 0.830 and 0.790, respectively, all higher than the original scale. The Cronbach’s α coefficient of all dimensions was greater than 0.8, indicating good internal consistency of the scale. The half-point reliability was 0.825>0.7, indicating high internal consistency of the scale. The retest reliability is 0.912, and reflection scale has good stability. In sumary, the MPSS has good reliability. To our knowledge, as one of the first study to validate the tool in different languages, the results of this study may be helpful for future research and clinical practice with maternal postpartum stress.

As we collected the questionnaire from the postpartum clinics, all selected women underwent postpartum check-ups 6 weeks after delivery. In terms of identity transition, most postpartum women exert all their energy into the child. As a result, they need more social and family support [44] and intimate relationship support [45]. As demand increases, parenting and postpartum fatigue increase, resulting in little attention to themselves. Moreover, most postpartum women choose to breastfeed. All the changes in hormone secretion and secretion levels [46] combined with the taboo on sex in China, jointly determine the proportion of stress, personal needs and fatigue, and infant nurturing. In contrast, the proportion of body changes and sexuality is higher. Personal needs and fatigue have the closest relationship with infant nurturing, while infant nurturing has the weakest association with body changes and sexuality.

Overall, the MPSS exhibited excellent internal consistency and significant inter-item and item-total correlations, indicating that the Chinese MPSS could confidently be used to assess maternal postpartum stress. The correlation between MPSS and EPDS, anxiety subscale in DASS-21and PSS-10 also supports the structural validity of MPSS. The results conform with previous research [19].

This study has the following limitations: Firstly, the subjects included only postpartum women in six hospitals in Nantong City. Hence, further research elsewhere is needed to validate Chinese versions of the MPSS. Secondly, since only those who meet certain requirements are selected, there are some deviations in the population selection in this study. Subsequent studies can measure postpartum women at different time points and verify the stress of postpartum women with multiple births, premature births, old age, and different marital statuses. In addition, this study is a cross-sectional study in which stress may exist by chance. Therefore, further cohort studies can be conducted to investigate maternal stress at different time points.

## Conclusions

In this study, the English version of MPSS was translated into Chinese. In addition, the reliability and validity test was carried out to form a Chinese version of MPSS containing 22 items in 3 subscales. The Chinese version of MPSS has good reliability and validity. After cultural adaptation, it has been used well among postpartum women in China. The MPSS is expected to provide a scientific basis for the early identification of postpartum women’s stress and formulation of early individualized intervention measures, which have certain clinical value and practical significance for postpartum women’s physical and mental health.

## Data Availability

All data produced in the present study are available upon reasonable request to the authors

## Abbreviations

MPSS: Maternal Postpartum Stress Scale
EFA: Exploratory factor analysis
CFA: Confirmatory factor analysis
EPDS: Edinburgh Postpartum Depression Scale
DASS-21: Depression, Anxiety, and Stress Scale-21
PSS-10: 10-item Perceived Stress Scale
S-CVI: Content validity index of the scale
HPSS: Hung Postpartum Stress Scale
I-CVI: Item Content Validity Index
IFI: Increment fitting index
TLI: Tucker Lewis index
CFI: Comparative fitting index

## Acknowledgements

The authors give thanks for all participants who volunteered to participate in this study.

## Authors’ contributions

Yanchi Wang completed the writing of this article. Qian Gao and Jin Liu played a key role in part of the data collection. Feng Zhang and Xujuan Xu put forward important revision suggestions and made corresponding modifications for this article. All authors contributed to the article and approved the submitted version.

## Funding

This work was supported by Social Science Foundation of Jiangsu Province (22SHB014).The funding sources had no role to play in the study design, the collection and interpretation of the data, writing of the report, or decision to submit this paper for publication.

### Availability of data and materials

The raw data of the current study would be available from the corresponding author on reasonable request.

### Declarations

#### Ethics approval and consent to participate

The study was reviewed and approved by the Ethics Committee of Affiliated Hospital of Nantong University (approval number: 2022-K50-01). All methods were performed in accordance with the relevant guidelines and regulations (Declaration of Helsinki). All participants have signed the Informed Consent.

### Consent for publication

Not Applicable.

### Competing interests

No potential conflicts of interest relevant to this article were reported.

## Notes

### Competing Interest Statement

The authors have declared no competing interest.

### Author Declarations

The study was reviewed and approved by the Ethics Committee of Affiliated Hospital of Nantong University (approval number: 2022-K50-01)

