## supplementary tables for "Maternal Postpartum Stress Scale: Translation and validation study of the Chinese version"

**Supplementary Table 1 The English version of the Maternal Postpartum Stress Scale**

| **items** | **Not at all**  (0) | **Slightly**  (1) | **Moderately** (2) | **Very** (3) | **Completely** (4 |
| --- | --- | --- | --- | --- | --- |
| 1. Choosing the appropriate way of feeding the baby (breastfeeding or formula) |  |  |  |  |  |
| 2. Baby’s irregular feeding pattern |  |  |  |  |  |
| 3. Insufficient milk supply when breastfeeding |  |  |  |  |  |
| 4. Baby’s development |  |  |  |  |  |
| 5. Baby’s health problems |  |  |  |  |  |
| 6. Recognizing the baby’s needs |  |  |  |  |  |
| 7. Impossibility to soothe a crying or upset baby |  |  |  |  |  |
| 8. Adjustment to frequent wake-ups |  |  |  |  |  |
| 9. Baby’s irregular patterns of daily sleep |  |  |  |  |  |
| 10. My fatigue and exhaustion |  |  |  |  |  |
| 11. The amount of the household chores |  |  |  |  |  |
| 12. Lack of help with the baby and household chores |  |  |  |  |  |
| 13. Being uncertain when to resume intercourse after childbirth |  |  |  |  |  |
| 14. Insufficient frequency of sexual intercourse |  |  |  |  |  |
| 15. Insufficient enjoyment in sexual intercourse |  |  |  |  |  |
| 16. The thought that my partner finds me unattractive |  |  |  |  |  |
| 17. The impossibility to return to the pre-pregnancy weight |  |  |  |  |  |
| 18. Lack of time for socializing with friends |  |  |  |  |  |
| 19. Lack of time for myself |  |  |  |  |  |
| 20. Physical appearance after childbirth |  |  |  |  |  |
| 21. Impossibility to complain to someone |  |  |  |  |  |
| 22. Loneliness at home with the baby |  |  |  |  |  |

**Supplementary Table 2 Item analysis (low-score groups=109, high-score groups=109)**

| Items | t | ΔMean | *P* value |
| --- | --- | --- | --- |
| A1 | 17.785 | 2.312 | <0.001 |
| A2 | 16.275 | 1.505 | <0.001 |
| A3 | 15.28 | 1.606 | <0.001 |
| A4 | 18.355 | 2.202 | <0.001 |
| A5 | 18.218 | 2.193 | <0.001 |
| A6 | 18.994 | 2.037 | <0.001 |
| A7 | 14.283 | 1.413 | <0.001 |
| A8 | 21.888 | 1.844 | <0.001 |
| A9 | 16.965 | 1.495 | <0.001 |
| A10 | 17.235 | 1.569 | <0.001 |
| A11 | 10.721 | 1.055 | <0.001 |
| A12 | 11.934 | 1.138 | <0.001 |
| A13 | 12.685 | 1.211 | <0.001 |
| A14 | 15.661 | 1.532 | <0.001 |
| A15 | 13.339 | 1.422 | <0.001 |
| A16 | 9.691 | 1.009 | <0.001 |
| A17 | 12.429 | 1.394 | <0.001 |
| A18 | 17.796 | 1.661 | <0.001 |
| A19 | 22.277 | 1.991 | <0.001 |
| A20 | 17.862 | 1.743 | <0.001 |
| A21 | 17.422 | 1.486 | <0.001 |
| A22 | 16.062 | 1.349 | <0.001 |

**Supplementary Table 3 Discretization trend among the Chinese version of MPSS**

|  |  | A1 | A2 | A3 | A4 | A5 | A6 | A7 | A8 | A9 | A10 | A11 |
| --- | --- | --- | --- | --- | --- | --- | --- | --- | --- | --- | --- | --- |
| Number of cases | effective | 406 | 406 | 406 | 406 | 406 | 406 | 406 | 406 | 406 | 406 | 406 |
|  | deletion | 0 | 0 | 0 | 0 | 0 | 0 | 0 | 0 | 0 | 0 | 0 |
| Average value | | 1.27 | 1.02 | 1.07 | 1.13 | 1.2 | 1.16 | 1.07 | 1.02 | 0.93 | 1.10 | 0.58 |
| Standard deviation | | 1.351 | 0.905 | 1.03 | 1.222 | 1.229 | 1.152 | 0.942 | 0.932 | 0.863 | 0.92 | 0.79 |
| Coefficient of Variation | | 1.0654 | 0.8877 | 0.964 | 1.0786 | 1.0222 | 0.9955 | 0.883 | 0.9095 | 0.9321 | 0.8357 | 1.353 |
|  |  | A12 | A13 | A14 | A15 | A16 | A17 | A18 | A19 | A20 | A21 | A22 |
| Number of cases | effective | 406 | 406 | 406 | 406 | 406 | 406 | 406 | 406 | 406 | 406 | 406 |
|  | deletion | 0 | 0 | 0 | 0 | 0 | 0 | 0 | 0 | 0 | 0 | 0 |
| Average value | | 0.62 | 0.60 | 0.76 | 0.62 | 0.41 | 0.82 | 0.89 | 1.13 | 0.95 | 0.80 | 0.76 |
| Standard deviation | | 0.834 | 0.831 | 0.933 | 0.929 | 0.764 | 0.984 | 0.91 | 0.997 | 1.001 | 0.816 | 0.768 |
| Coefficient of Variation | | 1.3493 | 1.3881 | 1.2295 | 1.4904 | 1.8675 | 1.1993 | 1.0204 | 0.8858 | 1.0557 | 1.0221 | 1.0156 |

**Supplementary Table 4** **Content validity of the questionnaire**

| Items | I-CVI | S-CVI |
| --- | --- | --- |
| A1 | 0.88 | 0.926 |
| A2 | 1 |  |
| A3 | 1 |  |
| A4 | 1 |  |
| A5 | 0.88 |  |
| A6 | 1 |  |
| A7 | 0.88 |  |
| A8 | 1 |  |
| A9 | 0.88 |  |
| A10 | 0.88 |  |
| A11 | 0.88 |  |
| A12 | 0.88 |  |
| A13 | 0.88 |  |
| A14 | 0.88 |  |
| A15 | 1 |  |
| A16 | 0.88 |  |
| A17 | 0.88 |  |
| A18 | 0.88 |  |
| A19 | 1 |  |
| A20 | 1 |  |
| A21 | 1 |  |
| A22 | 0.88 |  |

**Supplementary Table 5 The reliability of the MPSS**

| Variables | Cronbach’s α (n = 406) | | Test–retest  reliability  (n = 30) | half-test reliability |
| --- | --- | --- | --- | --- |
|  | English version | Chinese version |  |  |
| Total | 0.880 | 0.940 | 0.912 | 0.825 |
| personal needs and fatigue | 0.850 | 0.903 |  |  |
| infant nurturing | 0.830 | 0.911 |  |  |
| body changes and sexuality | 0.790 | 0.882 |  |  |

**Supplementary Table 6** **Correlation between each item of the MPSS and the total score (N = 406)**

| Items | Corrected  item-total  correlation | Cronbach alpha if the  item was deleted |
| --- | --- | --- |
| A1 | 0.631 | 0.938 |
| A2 | 0.642 | 0.937 |
| A3 | 0.608 | 0.937 |
| A4 | 0.700 | 0.936 |
| A5 | 0.698 | 0.936 |
| A6 | 0.684 | 0.936 |
| A7 | 0.550 | 0.938 |
| A8 | 0.722 | 0.936 |
| A9 | 0.643 | 0.937 |
| A10 | 0.613 | 0.937 |
| A11 | 0.492 | 0.939 |
| A12 | 0.530 | 0.938 |
| A13 | 0.576 | 0.938 |
| A14 | 0.633 | 0.937 |
| A15 | 0.596 | 0.937 |
| A16 | 0.526 | 0.938 |
| A17 | 0.525 | 0.939 |
| A18 | 0.685 | 0.936 |
| A19 | 0.756 | 0.935 |
| A20 | 0.666 | 0.936 |
| A21 | 0.672 | 0.937 |
| A22 | 0.677 | 0.937 |

**Supplementary Table 7 The evaluation of the goodness-of-fit of the model (n = 206)**

| χ2/df | CFI | IFI | TLI | RMSEA |
| --- | --- | --- | --- | --- |
| 2.167 | 0.918 | 0.919 | 0.907 | 0.075 |
